# Trends in Covid-19 risk-adjusted mortality rates in a single health system

**DOI:** 10.1101/2020.08.11.20172775

**Authors:** Leora I. Horwitz, Simon A. Jones, Robert J. Cerfolio, Fritz Francois, Joseph Greco, Bret Rudy, Christopher A. Petrilli

**Affiliations:** Center for Healthcare Innovation and Delivery Science, NYU Langone Health, New York, NY; Department of Population Health, NYU Grossman School of Medicine, New York, NY; Department of Medicine, NYU Grossman School of Medicine, New York, NY; Department of Surgery, NYU Grossman School of Medicine, New York, NY; NYU Winthrop Hospital, Mineola, NY; NYU Langone Hospital – Brooklyn, Brooklyn, NY

## Abstract

Early reports showed high mortality from Covid-19; by contrast, the current outbreaks in the southern and western United States are associated with fewer deaths, raising hope that treatments have improved. However, in Texas for instance, 63% of diagnosed cases are currently under 50, compared to only 52% nationally in March-April. Current demographics in Arizona and Florida are similar. Therefore, whether decreasing Covid-19 mortality rates are a reflection of changing demographics or represent improvements in clinical care is unknown. We assessed outcomes over time in a single health system, accounting for changes in demographics and clinical factors.

Methods

We analyzed biweekly mortality rates for admissions between March 1 and June 20, 2020 in a single health system in New York City. Outcomes were obtained as of July 14, 2020. We included all hospitalizations with laboratory-confirmed Covid-19 disease. Patients with multiple hospitalizations (N=157, 3.3%) were included repeatedly if they continued to have laboratory-confirmed disease. Mortality was defined as in-hospital death or discharge to hospice care. Based on prior literature, we constructed a multivariable logistic regression model to generate expected risk of death, adjusting for age; sex; self-reported race and ethnicity; body mass index; smoking history; presence of hypertension, heart failure, hyperlipidemia, coronary artery disease, diabetes, cancer, chronic kidney disease, or pulmonary disease individually as dummy variables; and admission oxygen saturation, D-dimer, C reactive protein, ferritin, and cycle threshold for RNA detection. All data were obtained from the electronic health record. We then calculated the sum of observed and expected deaths in each two-week period and multiplied each period's observed/expected (O/E) risk by the overall average crude mortality to generate biweekly adjusted rates. We calculated Poisson control limits and indicated points outside the control limits as significantly different, following statistical process control standards. The NYU institutional review board approved the study and granted a waiver of consent.

Results

We included 4,689 hospitalizations, of which 4,661 (99.4%) had died or been discharged. The median age, and the proportion male or with any comorbidity decreased over time; median real-time PCR cycle threshold increased (indicating relatively less concentration of virus) (Table). For instance, median age decreased from 67 years in the first two weeks to 49 in the last two. Peak hospitalizations were during the fifth and sixth study weeks, which accounted for 40% of the hospitalizations. Median length of stay for patients who died or were discharged to hospice was 8 days (interquartile range, 4-16).

Unadjusted mortality dropped each period, from 30.2% in the first two weeks to 3% in the last two weeks, with the last eight weeks being lower than the 95% control limits. Risk adjustment partially attenuated the mortality decline, but adjusted mortality rates in the second-to-last two weeks remained outside the control limits (Figure, Table). The O/E risk of mortality decreased from 1.07 (0.64-1.67) in the first two weeks to 0.39 (0.08-1.12) in the last two weeks.

Discussion

In this 16-week study of Covid-19 mortality at a single health system, we found that changes in demographics and severity of illness at presentation account for some, but not all, of the decrease in unadjusted mortality. Even after risk adjustment for a variety of clinical and demographic factors, mortality was significantly lower towards the end of the study period.

Incremental improvements in outcomes are likely a combination of increasing clinical experience, decreasing hospital volume, growing use of new pharmacologic treatments (such as corticosteroids, remdesivir and anti-cytokine treatments), non-pharmacologic treatments (such as proning), earlier intervention, community awareness, and lower viral load exposure from increasing mask wearing and social distancing. It is also possible that earlier periods had a more virulent circulating strain.

In summary, data from one health system suggest that Covid-19 remains a serious disease for high risk patients, but that outcomes may be improving.

## Methods

We analyzed biweekly mortality rates for admissions between March 1 and June 20, 2020 in a single health system in New York City. Outcomes were obtained as of July 14, 2020. We included all hospitalizations with laboratory-confirmed Covid-19 disease. Patients with multiple hospitalizations (N=157, 3.3%) were included repeatedly if they continued to have laboratory-confirmed disease. Mortality was defined as in-hospital death or discharge to hospice care. Based on prior literature, we constructed a multivariable logistic regression model to generate expected risk of death, adjusting for age; sex; self-reported race and ethnicity; body mass index; smoking history; presence of hypertension, heart failure, hyperlipidemia, coronary artery disease, diabetes, cancer, chronic kidney disease, or pulmonary disease individually as dummy variables; and admission oxygen saturation, D-dimer, C reactive protein, ferritin, and cycle threshold for RNA detection.^3^ All data were obtained from the electronic health record. We then calculated the sum of observed and expected deaths in each two-week period and multiplied each period’s observed/expected (O/E) risk by the overall average crude mortality to generate biweekly adjusted rates. We calculated Poisson control limits and indicated points outside the control limits as significantly different, following statistical process control standards.^4^ The NYU institutional review board approved the study and granted a waiver of consent.

## Results

**Figure:**
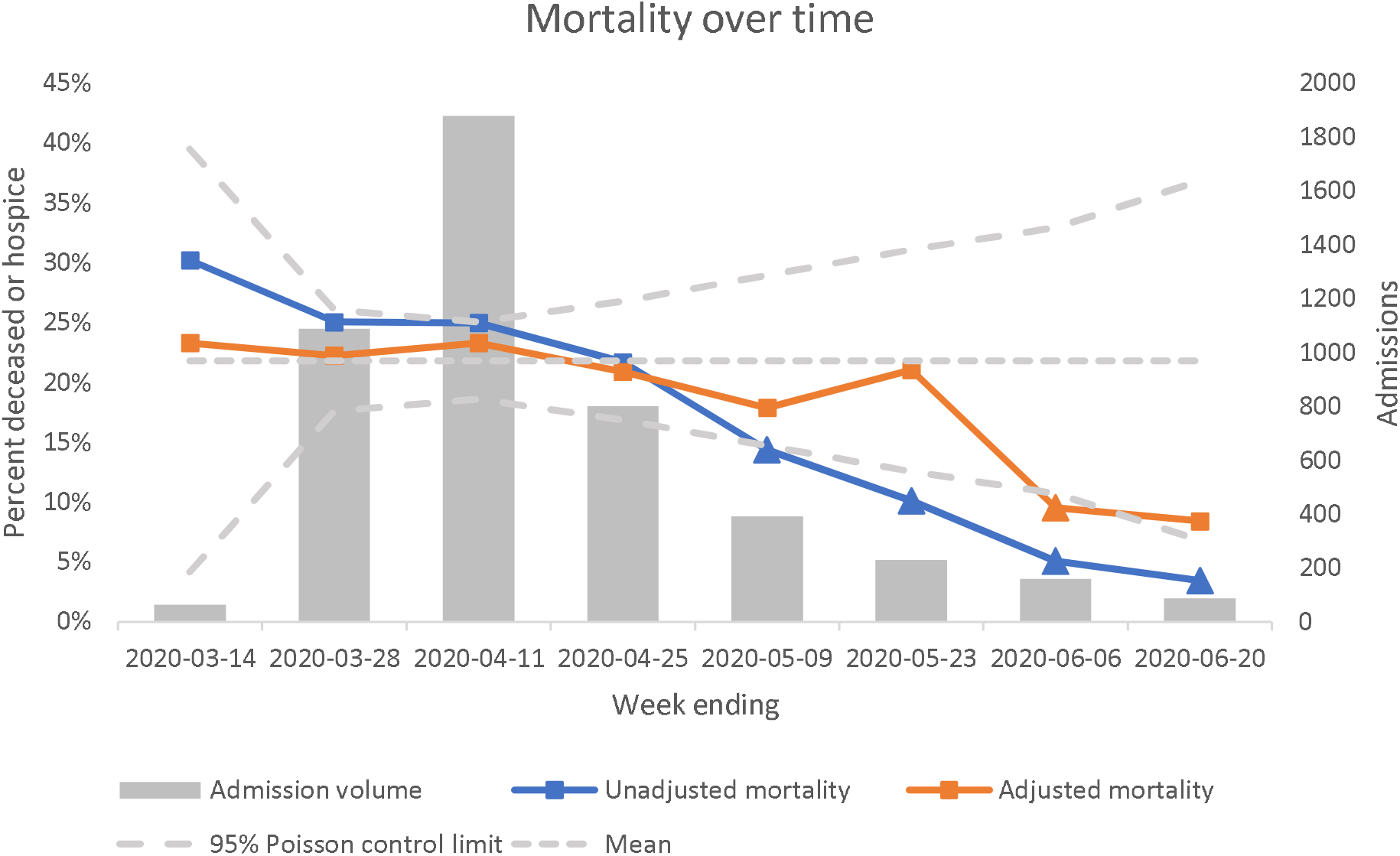
Biweekly mortality over time. Triangular markers indicate points outside control limits.

**Table:**
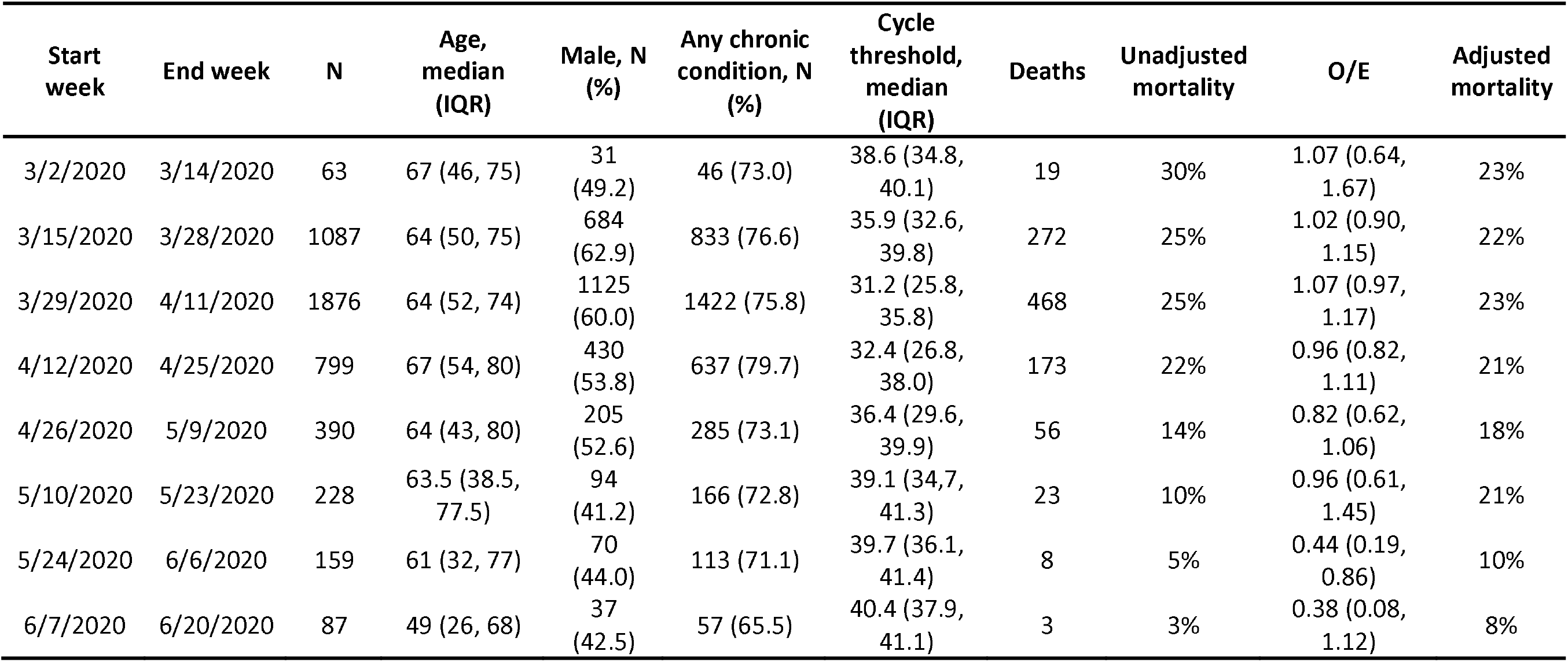
Demographics and mortality by week of admission

## Discussion

Incremental improvements in outcomes are likely a combination of increasing clinical experience, decreasing hospital volume, growing use of new pharmacologic treatments (such as corticosteroids,^5^ remdesivir^6^ and anti-cytokine treatments), non-pharmacologic treatments (such as proning), earlier intervention, community awareness, and lower viral load exposure from increasing mask wearing and social distancing. It is also possible that earlier periods had a more virulent circulating strain.high risk patients, but that outcomes may be improving.

## Data Availability

Data for this study are not publicly available.

## To the Editor

Early reports showed high mortality from Covid-19; by contrast, the current outbreaks in the southern and western United States are associated with fewer deaths, raising hope that treatments have improved. However, in Texas for instance, 63% of diagnosed cases are currently under 50,^1^ compared to only 52% nationally in March-April.^2^ Current demographics in Arizona and Florida are similar. Therefore, whether decreasing Covid-19 mortality rates are a reflection of changing demographics or represent improvements in clinical care is unknown. We assessed outcomes overtime in a single health system, accounting for changes in demographics and clinical factors.

## References

1. Texas Department of State Health Services. Texas COVID-19 Data. 2020; https://www.dshs.texas.gov/coronavirus/additionaldata/. Accessed 22 July, 2020.

2. Centers for Disease Control and Prevention. COVIDView. 2020; https://www.cdc.gov/coronavirus/2019-ncov/covid-data/covidview/index.html. Accessed 22 July, 2020.

3. Petrilli CM, Jones SA, Yang J, et al. Factors associated with hospital admission and critical illness among 5279 people with coronavirus disease 2019 in New York City: prospective cohort study. Bmj. 2020;369:ml966.

4. Spiegelhalter DJ. Funnel plots for comparing institutional performance. Stat Med. 2005;24(8):1185-1202.

5. Recovery Collaborative Group, Horby P, Lim WS, et al. Dexamethasone in Hospitalized Patients with Covid-19 - Preliminary Report. N Engl J Med. 2020.

6. Beigel JH, Tomashek KM, Dodd LE, et al. Remdesivir for the Treatment of Covid-19 - Preliminary Report. N Engl J Med. 2020.

